# Integrating Clinical Research with Electronic Health Records to Improve Diversity in Research: Findings from an Urban Hospital System

**DOI:** 10.1101/2024.09.18.24313820

**Authors:** Edward H. Brown, Nicholas C. Peiper, Stephen Furmanek, Kelly McCants

**Affiliations:** Population Health Research, Norton Healthcare, Louisville, Kentucky, USA; Institute for Health Equity, Norton Healthcare, Louisville, Kentucky, USA; Department of Epidemiology and Population Health, University of Louisville, Louisville, Kentucky, USA; Department of Psychological and Brain Sciences, University of Louisville, Louisville, Kentucky, USA; Advanced Heart Failure and Recovery Program, Norton Heart and Vascular Institute, Louisville, Kentucky, USA

## Abstract

**Background:** The under-representation of racial, ethnic, and other minority populations in clinical research has threatened the validity of novel therapeutic interventions and exacerbated the longstanding inequities in health outcomes. Despite attention and mandates across institutions and federal agencies to address these disparities, effective and actionable strategies have remained a subject of debate in the existing literature. Thus, the need for comprehensive and rigorous methods to assess diversity in study population samples as well as strategies for improved recruitment and retention has been made clear.

**Objective:** Examine the distribution of research participant demographics at Norton Healthcare (NHC) and compare to applicable benchmarks from the overall NHC patient population and local census.

**Design:** Successive-independent samples comparison of EPIC electronic health records (EHR) and census data from Jefferson County.

**Participants:** A total of 2,053 adult research participants enrolled at NHC from 2020 – 2024.

**Main Measures:** Demographic data were reported as frequency and percentage across the three benchmarks. The z-test for independent proportions was used to compare the research participant demographics to the NHC patients and Jefferson County during a consistent time period. Temporal trends in research participant demographics were also examined.

**Key Results:** Apart from being relatively older, the NHC research participant population closely resembled the NHC patient population. A similar age-related pattern emerged as well as an over-representation of White individuals in the research participant group when compared to the census data. When looking at research participant demographic trends overtime, increases in White, Black and older cohorts were noted while decreases were observed in Hispanic/Latinx and younger cohorts. Trends related to participant sex remained stable.

**Conclusions:** The findings from this study will inform future strategies for setting enrollment goals and facilitate the creation of tools and metrics to evaluate appropriate standards for diversity in clinical research study population samples.

## Introduction

Historically, clinical research studies have lacked equitable inclusion of people from racial and ethnic minority (REM) populations.[1] As a result of inequitable inclusion of REM populations, the purported effectiveness of treatments, interventions, and other novel therapeutics may not generalize beyond White populations and other commonly favored groups. Moreover, the underrepresentation of REM populations in clinical research studies may preclude potential benefits to morbidity, mortality, and health-related quality of life.[2,3] Underrepresentation may also exacerbate long-standing structural factors (e.g., medical distrust, structural racism) stemming from exploitation of REM populations by the United States (US) medical system.[4,5] In addition, underrepresentation is associated with extremely high financial and social costs in the range of hundreds of billions of dollars.[6,7]

Addressing underrepresentation of REM populations in clinical research studies has been a longstanding goal of the US Department of Health and Human Services (HHS). For example, the National Institutes of Health (NIH) Revitalization Act of 1993 mandated appropriate inclusion of REM populations and women in clinical trials.[8] Additionally, this policy required clinical research sponsored by the NIH to address the inclusion of historically marginalized and medically underserved demographic groups in grant applications as well as Phase 3 clinical trials. Despite the promulgation of a variety of HHS policies focusing on improving representation in clinical research studies, inclusion of REM populations has remained unacceptably low over the past 30 years.[8,9]

More recently, the National Academies of Science, Engineering, and Medicine (NASEM) published a comprehensive report entitled *Improving Representation in Clinical Trials and Research*.[10] The report reiterated the history and current trends in underrepresentation, specifically calling urgent attention to 17 recommendations focused on improving investment, infrastructure, transparency, and accountability across the entire biomedical enterprise. Similarly, the Food and Drug Administration (FDA) recently released guidance documents to improve enrollment of REM populations, including an impending policy that will require researchers and companies seeking approval for late-stage clinical trials to submit a diversity enrollment plan.[11] Given the calls from HHS, NASEM, and other medical governing bodies, it is imperative that healthcare systems rigorously evaluate the representation of REM populations in clinical research studies and formulate culturally appropriate strategies to increase enrollment diversity.[10–12]

In response to this imperative, we present findings from an ongoing clinical research program to increase representation in clinical trials and research within a large, urban healthcare system. The primary objective of this project is to examine the demographic composition of research participants and establish preliminary benchmarks for setting equitable enrollment goals in future studies. We detail our design for integrating research data with electronic health records (EHR) to evaluate trends in enrollment from 2020 to 2024 across REM populations and benchmark the demographic composition of research participants against the overall patient population and community Census. We highlight potential benefits of utilizing research data to set enrollment benchmarks for REMs and other health disparity populations. We also discuss limitations of the relevant data sources, along with the need for community engagement and cross-sector partnerships to address systemic issues driving underrepresentation.

## Methods

### Study Context

The Norton Healthcare (NHC) Institute for Health Equity has an ongoing project that began in January 2023 to improve diversity in clinical research. This study represents the data-driven component of the project to integrate clinical research data with EHR’s and generate benchmarks for quality improvement activities. The other component involves a workforce development program in partnership with historically black college and universities (HBCU), state and local universities, and minority-serving institutions focused on creating sustainable pipelines for diverse populations to enter the clinical research workforce.

NHC is the largest healthcare system in Louisville, Kentucky, with over 2 million patient interactions per year. NHC has five Louisville-based hospitals and three in Southern Indiana. There are 500 newly initiated clinical trials conducted across NHC per year, covering a wide variety of medical conditions. Louisville is Kentucky’s largest city with a population of around 625,000 residents, while Louisville Metro includes the city along with 12 surrounding counties in both KY and IN for a total of 1.4 million people. While Louisville is experiencing rapid economic growth, structural racism persists through unequal distributions of quality healthcare, police violence, geographic isolation, equitable allocation of city resources, and limited job opportunities.[13–19] For example, life expectancy in the Northwest Core of Louisville is over 15 years less than the Eastern Core.[20] Given the longstanding racial injustices in Louisville, this project was designed with an emphasis on social determinants of health (SDOH) and community partnerships.

### Study Sample

Participants included adults at least 18 years old who were enrolled into a clinical research study from January 2020 to March 2024. Research participant data were matched to EHR data to obtain demographic data. We compared the demographic distribution of research participants to the NHC adult patient population and Louisville adult population. Patient data were derived from EHR summary reports using Microsoft SQL Server Management Studio and population census data were based upon the American Community Survey 2022 5-year estimates for Jefferson County, KY.

### Study Measures

We examined race (White, Black, Asian, Native Hawaiian and Pacific Islander [NH/PI], American Indian and Alaska Native [AI/AN], multiracial, and other), ethnicity (Hispanic), biological sex assigned at birth, and age (18-24, 25-34, 35-44, 45-54, 55-64, 65-74, 75-84, 85+). Demographics were classified according to US Census definitions. Year of enrollment included 2020, 2021, 2022, 2023, and 2024 (only Q1).

### Statistical Analysis

Participant demographic data were reported as frequency and percentage. The z-test for independent proportions was used to compare the research participant demographics to the NHC patients and Jefferson County demographics. P-values less than 0.05 were used to define statistically significant differences between populations. Cohen’s *h* was used to calculate effect size for the differences.[21] Each subcategory was evaluated for statistical significance (*p*-value) and effect size (Cohen’s *h*), achieving practical relevance in this analysis by meeting the minimum criteria of *p ≤* 0.05 and *h ≥* 0.2.[22] As a sensitivity analysis, we conducted stratified analyses by year of enrollment to explore time trends in demographic enrollment.

## Results

This study examined the demographic distribution of adult research participants from January 2020 – March 2024 (n=2,053) and compared those results with both the NHC patient population (2020 – 2024) and local census data from Jefferson County (Louisville, KY) via the American Community Survey 2022 5-year estimates (Table 1). When comparing the research participant group to the overall patient population, the 25-34 age group represented a significantly smaller (5.2% vs. 11.8%, Cohen’s h=0.24) and the 65-74 age group (28.8% vs. 19.5%, Cohen’s h=0.22) represented a significantly larger proportion. Next, when comparing the research participant group to local census, several differences were noted according to race and age. Regarding race, the research participant group represented a significantly greater proportion of White people (79.5% vs. 70.9%, Cohen’s h=0.2). Regarding age, the research participant group represented a significantly smaller proportion of ages 18-24 (3.6% vs. 11.1%, Cohen’s h=0.3) and 25-34 (5.2% vs. 18.7%, Cohen’s h=0.43). Significantly larger proportions were observed among the research participants for ages 65-74 (28.8% vs. 12.9%, Cohen’s h=0.4) and 75-84 (14.6% vs. 6.0%, Cohen’s h=0.29).

**Table 1.** Demographic distribution of participants enrolled in clinical research studies, 2020-2024*.

| Demographics | Enrolled<br>Participants<br>2020-2024<br>n=2,053 | NHC<br>Patients<br>2020-2024<br>n=2,362,097 | Effect<br>Size | p-value | Jefferson<br>County<br>population*<br>n=608,210 | Effect<br>Size | p-value |
| --- | --- | --- | --- | --- | --- | --- | --- |
| **Race** |  |  |  |  |  |  |  |
| White | 1633 (79.5%) | 76.6% | 0.07 | 0.002 | 70.9% | **0.20** | **<0.001** |
| Black or African American | 298 (14.5%) | 16.9% | 0.07 | 0.004 | 20.4% | 0.16 | **<0.001** |
| American Indian or Alaskan Native | - | - | - | - | - | - | - |
| Asian | 20 (1.0%) | 1.4% | 0.04 | 0.122 | 0.8% | 0.14 | **<0.001** |
| Native Hawaiian or Pacific Islander | - | - | - | - | <0.1% | - | - |
| Other Race | 65 (3.2%) | 3.7% | 0.03 | 0.221 | 5.7% | 0.12 | **<0.001** |
| Unknown Race | 32 (1.6%) | 2.3% | 0.05 | 0.030 | 0.0% | - | - |
| **Ethnicity** |  |  |  |  |  |  |  |
| Hispanic or Latinx | 53 (2.6%) | 3.3% | 0.04 | 0.078 | 5.3% | 0.14 | **<0.001** |
| **Biological Sex** |  |  |  |  |  |  |  |
| Female | 1217 (59.2%) | 65.0% | 0.12 | **<0.001** | 51.3% | 0.16 | **<0.001** |
| **Age** |  |  |  |  |  |  |  |
| 18 – 24 | 73 (3.6%) | 6.4% | 0.13 | **<0.001** | 11.1% | **0.30** | **<0.001** |
| 25 – 34 | 107 (5.2%) | 11.8% | **0.24** | **<0.001** | 18.7% | **0.43** | **<0.001** |
| 35 – 44 | 249 (12.1%) | 12.9% | 0.02 | 0.312 | 16.4% | 0.12 | **<0.001** |
| 45 – 54 | 262 (12.8%) | 14.5% | 0.05 | 0.027 | 15.4% | 0.07 | 0.001 |
| 55 – 64 | 412 (20.1%) | 18.3% | 0.05 | 0.041 | 16.9% | 0.08 | **<0.001** |
| 65 – 74 | 592 (28.8%) | 19.5% | **0.22** | **<0.001** | 12.9% | **0.40** | **<0.001** |
| 75 – 84 | 299 (14.6%) | 12.3% | 0.07 | 0.002 | 6.0% | **0.29** | **<0.001** |
| 85 and older | 59 (2.9%) | 4.2% | 0.07 | 0.003 | 2.5% | 0.02 | 0.310 |

\*Includes participants enrolled in a clinical research study from January 1, 2020 to March 31, 2024. Cell sizes below 10 are suppressed. Bolded values denote practically relevant findings meeting the criteria of $p \leq 0.05$ and Cohen's $h \geq 0.2$ .
Abbreviations: NHC, Norton Healthcare

In addition to comparing demographic distributions to other benchmarks, we examined annual trends across each demographic subpopulation. The proportion of White research participation (77.4% in 2020 to 81.2% in 2024) and Black participation (13.9% in 2020 to 14.8% in 2024) slightly increased, while all other groups seemed to trend lower or remained stable across the time frame examined (Figure 1). The proportion of Hispanic/Latinx participation trended downward (4.2% in 2020 to 1.3% in 2024) while female participation remained stable (Figure 2, 3). For age (Figure 4), the research participant population sample skewed towards the older age groups across time (28.8% of 65 and older in 2020 compared to 85.2% in 2024).

**Figure 1.**
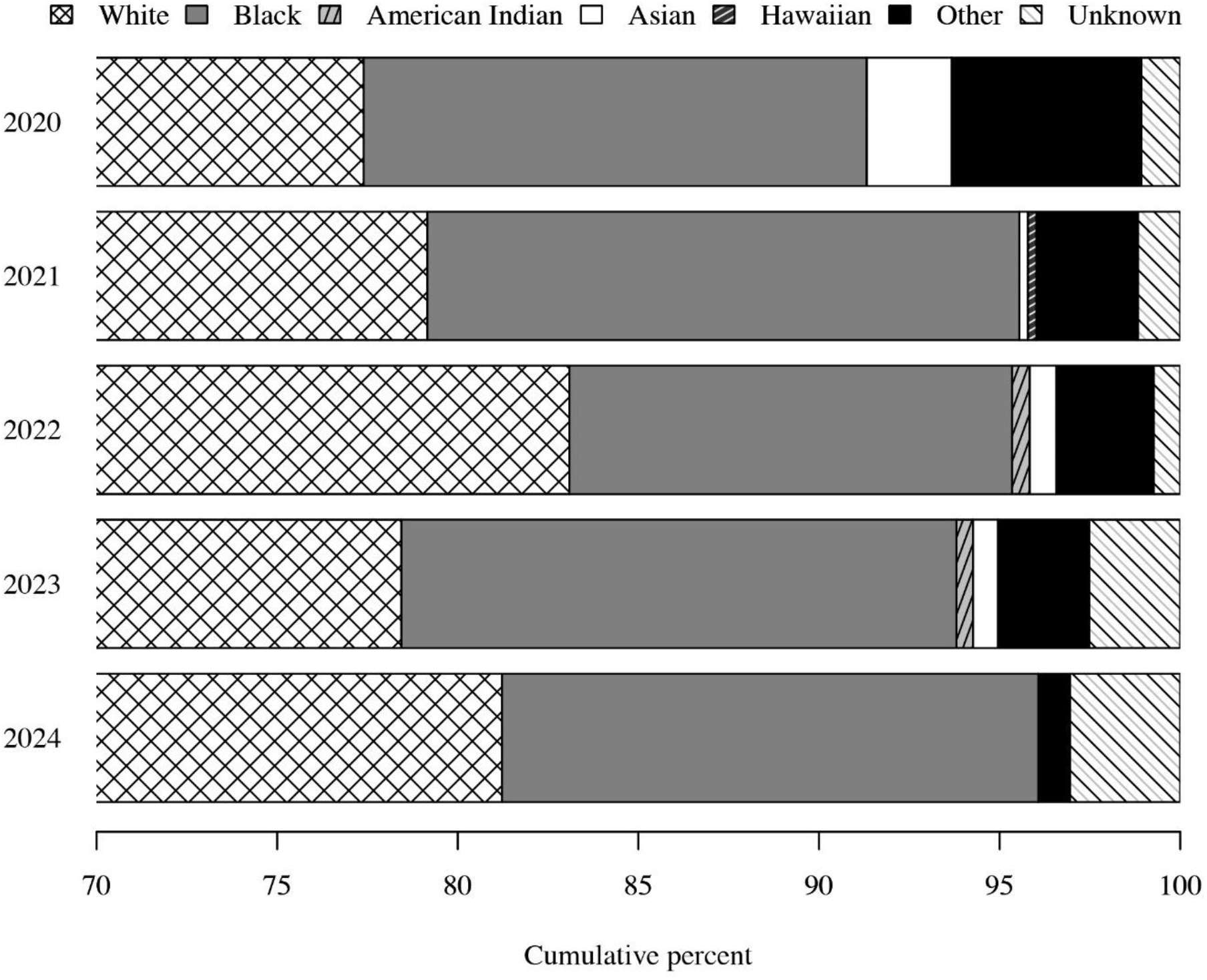
Trends in the distribution of race among participants enrolled in clinical research studies, 2020-2024* *Includes participants enrolled in a clinical research study from January 1, 2020 to March 31, 2024. The y-axis was scaled to facilitate interpretation of small proportions.

**Figure 2.**
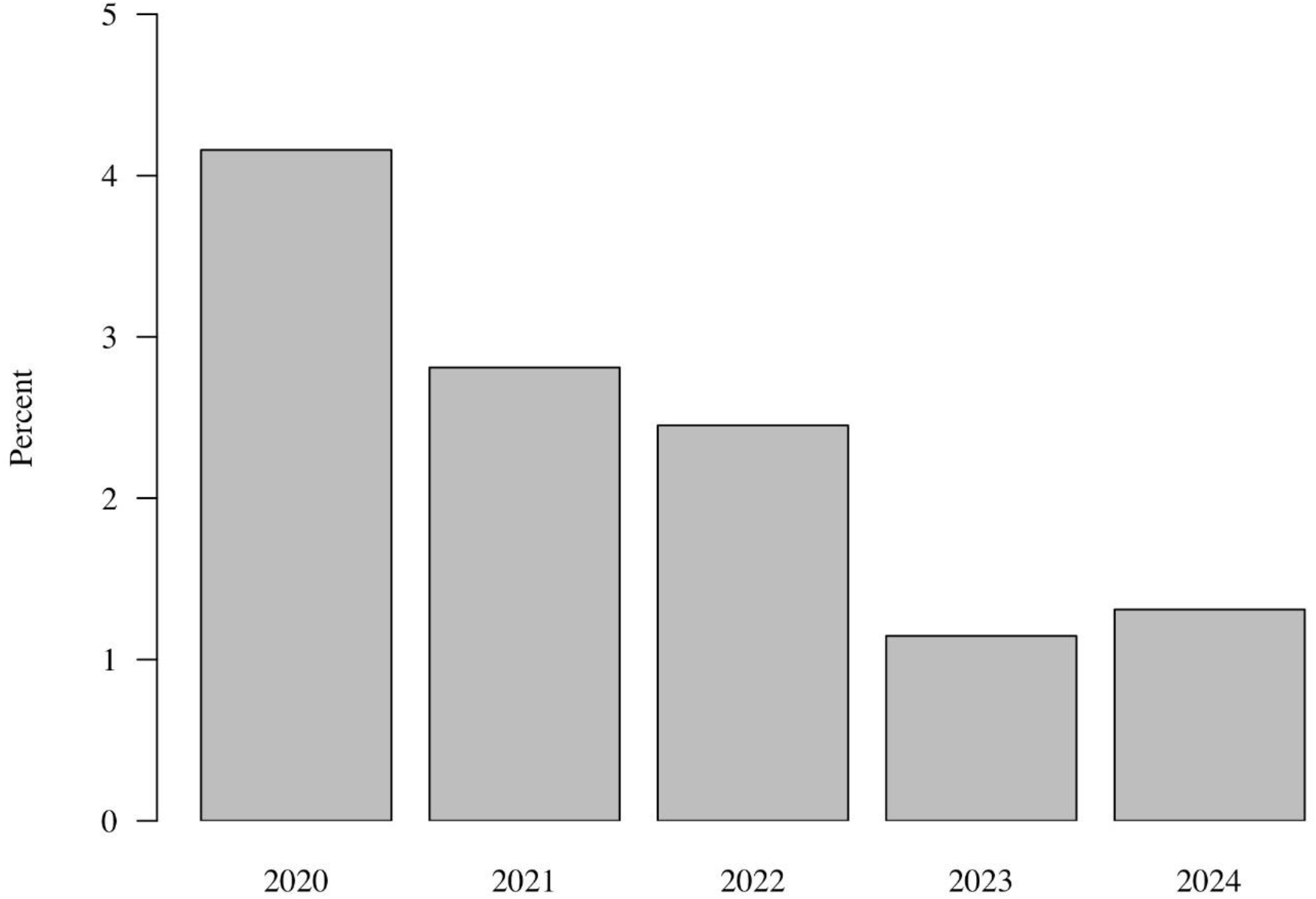
Trends in the distribution of Hispanic participants enrolled in clinical research studies, 2020-2024* *Includes participants enrolled in a clinical research study from January 1, 2020 to March 31, 2024. The y-axis was scaled to facilitate interpretation of small proportions.

**Figure 3.**
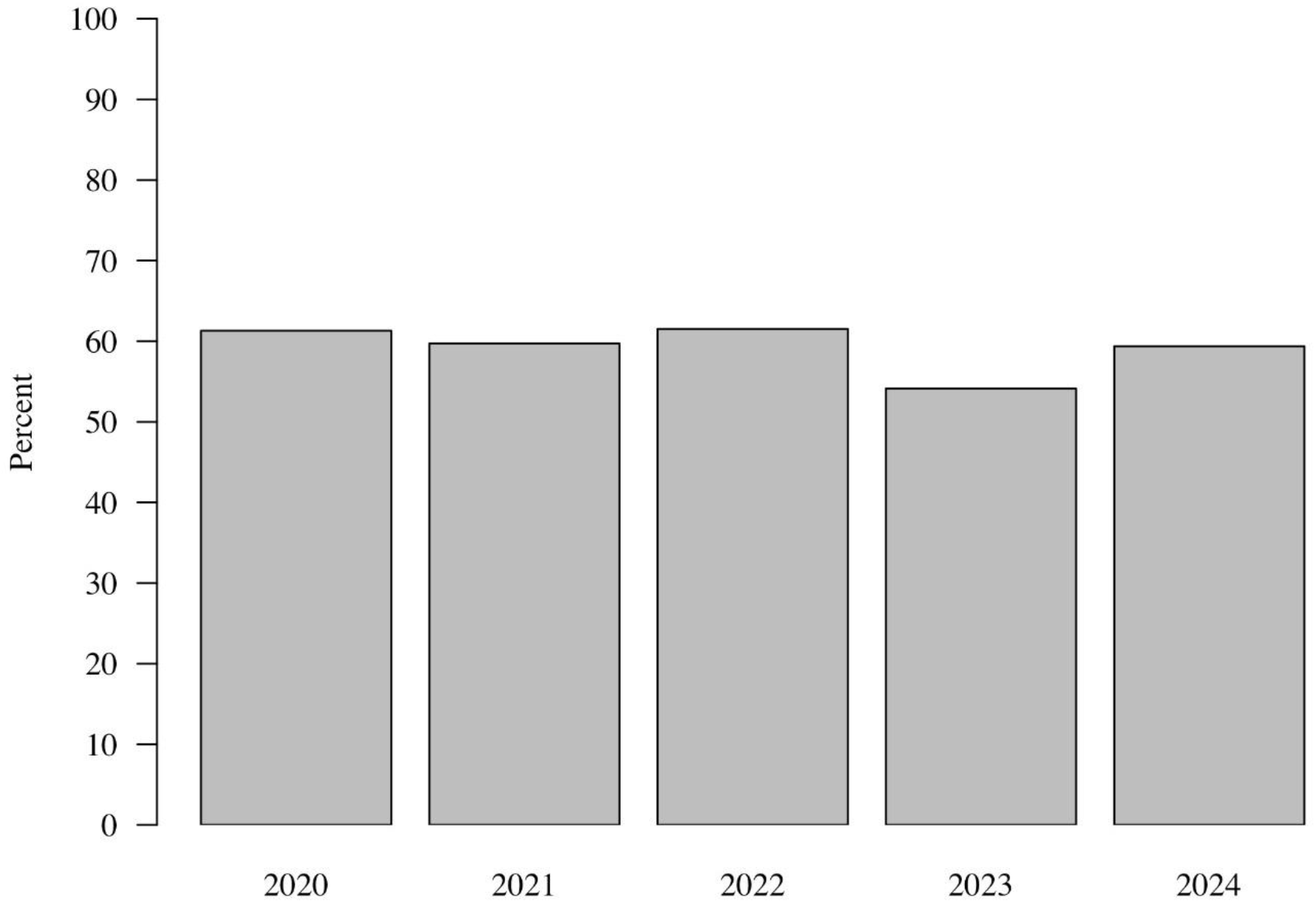
Trends in the distribution of biological females among participants enrolled in clinical research studies, 2020-2024* *Includes participants enrolled in a clinical research study from January 1, 2020 to March 31, 2024

**Figure 4.**
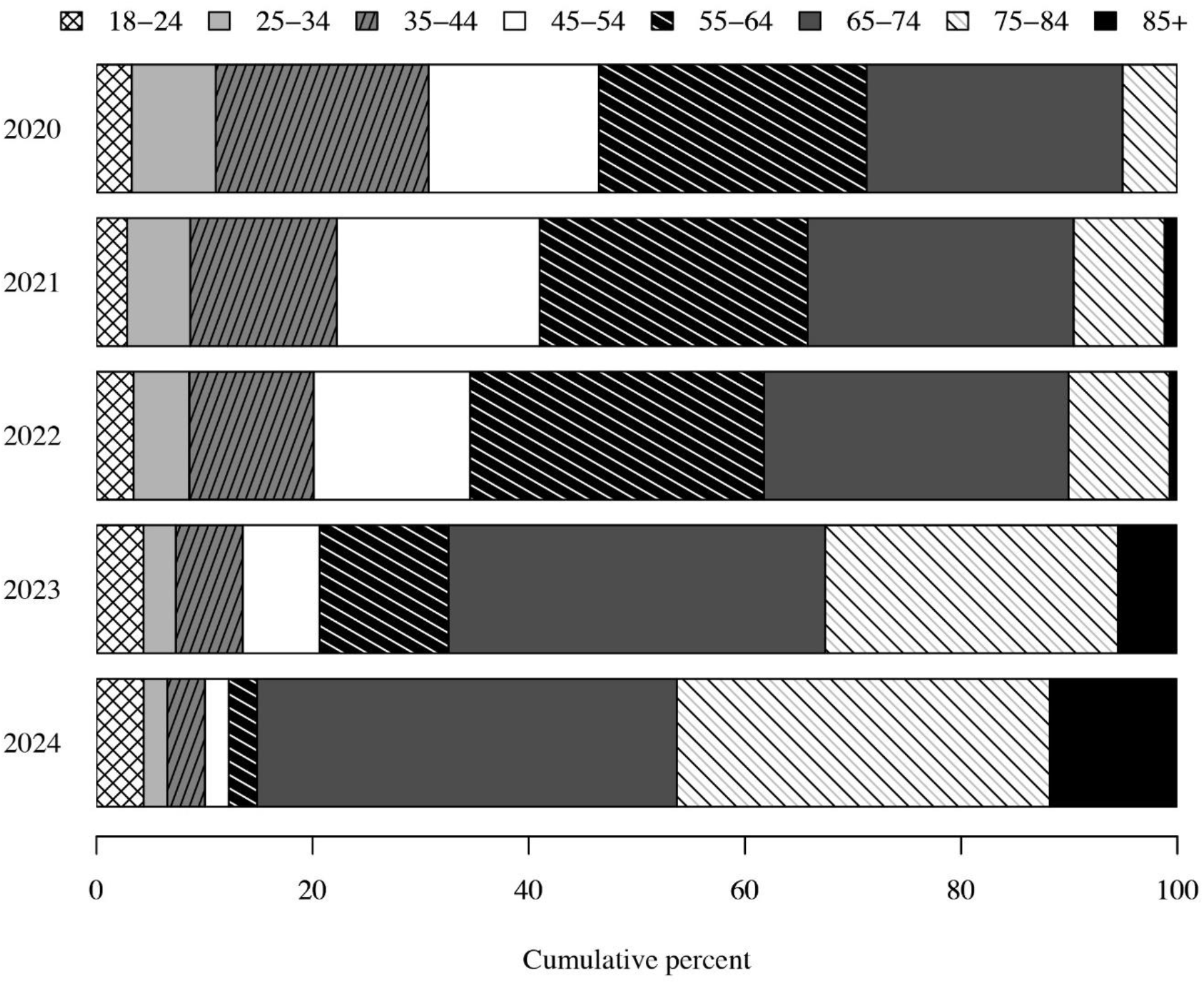
Trends in the age distribution of participants enrolled in clinical research studies, 2020-2024* *Includes participants enrolled in a clinical research study from January 1, 2020 to March 31, 2024

## Discussion

This study detailed findings to increase representation in clinical trials and research within an urban hospital system. Study enrollment data were synthesized from a research platform and integrated with EHRs to evaluate trends in enrollment from 2020 to 2024 across REM populations and benchmark the demographic composition to the overall patient population and Census. Findings from this study represent a crucial first step in developing equitable enrollment goals and evaluation metrics in future studies both at the aggregate and study-level.[10]

Our initial examination included all research participants enrolled across studies from 2020 to 2024. Apart from being older, we found the research population matches the overall patient population, including REMs. While this suggests that the distribution of REMs broadly mirror patterns in the larger patient population, emergent frameworks suggest setting enrollment goals for REMs that are at least at the Census level.[23] Comparisons to real-world patient data and the scientific literature may further determine whether certain enrollment thresholds should be higher than Census estimates. In this study, we found higher proportions of REMs in the local Census estimates, which is highly consistent with the extant literature. We observed fluctuations in the distribution of certain REM participants from 2020 to 2024 (e.g., increases in Blacks, decreases in Hispanics) and increases in participant age, while the proportion of females remained steady. These trends may have been a function of research studies covering specific disease states with varying frequency and severity among REMs and other demographic subpopulations.[9,10,23] As findings from this study provide preliminary enrollment benchmarks at the aggregate level, additional studies that examine disease-specific trends will be necessary to develop enrollment thresholds with greater granularity by disease status and study type (e.g., intervention vs. non-intervention).[23–25]

Another key finding was that research participant data were readily linked to EHRs and compared to patient- and community-level trends. We demonstrated the utility of evaluating enrollment data to develop practical metrics for clinical research participation, although establishing equitable thresholds for follow-up and completion of studies also warrants attention. For example, adult and pediatric studies demonstrate that REMs tend to have higher rates of attrition in clinical trials.[26] Other studies have found inequities in attrition based upon geography, socioeconomic status, gender identity, and sexual orientation. Targeted evaluations will be conducted to ensure equitable follow-up and improved assessment of study outcomes. Findings from this study also align with emergent guidelines for increased research diversity. For example, FDA recently released draft guidance on June 26, 2024 about Diversity Action Plans and requirements for sponsors.[11] Similarly, the US Office of Management and Budget announced in March 2024 changes to how race and ethnicity will be collected to reduce misclassification.[27] Given the evolving policy landscape and widespread implications for improving diversity in clinical research, SDOH driving poor enrollment and follow-up among REMs will require more explicit emphasis in the development of quality improvement programs.[2]

A major component of understanding SDOH will include participatory action research (PAR) with the Louisville community.[28] As part of this effort, an advisory board of community experts from REM backgrounds has been assembled to provide input on systematically gathering community perspectives on the impact of SDOH on clinical trial participation. The board ranges from scientific experts in academia and industry to directors of social and health organizations in Louisville. Utilization of this board will help facilitate better access to REMs and other vulnerable populations. Our potential reach into these communities is amplified by the marked increases in racial and ethnic diversity in Louisville over the past 30 years, although persistent racial segregation continues to pose significant barriers. Given the complex social and historical context of racial inequities in Louisville, qualitative PAR studies have been designed to understand community perspectives on clinical research, such as strategies to rebuild trust and link REMs to active studies.[29,30] These perspectives will coincide with the opening of a new NHC hospital in West Louisville, which will be co-located with a variety of social service agencies (e.g., Goodwill, Volunteers of America). As such, these PAR and other community-engaged studies will provide key insights on how to increase clinical research diversity in partnership with social service agencies and community-based organizations.[31–33]

Several limitations to this study are acknowledged. Our study sample introduces a selection bias given that all research participants studied were only enrolled within a single healthcare system and not across the whole locality. The sample was also not stratified by disease state, limiting the generalizability of the study to participants overall. Follow-up studies are underway to further delineate representation across disease states and study types (e.g., intervention, non-intervention). Additionally, we only examined enrollment data and did not examine demographic distributions across the whole research cascade (e.g., withdrawal, follow-up, completion). The integrated data from the EHR only included enrollment status by year, precluding our ability to examine time trends with more granularity (e.g., quarterly, monthly). The timeframe of this study included the initial phase of the COVID-19 pandemic, which may have impacted enrollment and inclusion. Lastly, a Type 1 error cannot be ruled out with the significant differences between the patient and census data, although our effect size estimates helped identify practically significant differences. Additional studies are needed to examine other measures of practical significance and their application to examining how the distribution of demographic subpopulations compare to relevant benchmarks.

This study examined clinical research enrollment data to establish preliminary benchmarks for setting equitable enrollment goals in future quality improvement programs. Follow-up studies are now necessary to investigate differences by disease status and study type. PAR studies will incorporate the perspectives and lived experiences of community members into future programmatic efforts. Similarly, studies of clinical research staff will be necessary to better understand measurement of participant demographics and SDOH.[34–36] Taken together, this study provides foundational information for developing data-driven tools for use by researchers to set enrollment goals and align with the US federal government’s recent strategies to improve enrollment of REM populations.

## Data Availability

All data produced in the present work are contained in the manuscript.

## Acknowledgments

This study was supported by an unrestricted grant through the Abbott Laboratories Diversity in Clinical Research Initiative. Dr. Peiper reports research support through the National Institutes of Health and Centers for Disease Control and Prevention (CDC). Dr. Peiper reports scientific advisory fees and stock options from Meru Health. Mr. Brown reports consultation fees from Meru Health and research support through the CDA Foundation.

